# The role of repeat training in participants undertaking take home naloxone interventions

**DOI:** 10.1101/2020.04.26.20074542

**Authors:** Maria Bravo, Lauren Monds, Llewellyn Mills, Phillip Read, Mary Ellen Harrod, Suzanne Nielsen, Marianne Jauncey, Consuelo Rivas, Annie Malcolm, Rosie Gilliver, Nicholas Lintzeris

**Affiliations:** Drug and Alcohol Services, South Eastern Sydney Local Health District (SESLHD), Sydney, Australia; Discipline of Addiction Medicine, Central Clinical School, The University of Sydney, Sydney, Australia; NSW Drug and Alcohol Clinical Research and Improvement Network (DACRIN), Sydney, Australia; Kirketon Road Centre, SESLHD, Sydney, Australia; NSW Users and AIDS Association (NUAA), Sydney, Australia; Monash Addiction Research Centre, Monash University, Melbourne, Australia; Uniting Sydney Medically Supervised Injecting Centre

**Keywords:** opioid overdose, take home naloxone, brief intervention, training evaluation

## Abstract

**Introduction and Aims:** Training of consumers is seen as a necessary component of take home naloxone (THN) interventions, with demonstrated improvements in consumer knowledge, attitudes and self-efficacy. However, we query whether re-training is necessary component for individuals renewing naloxone supplies who have previously completed a THN intervention.

**Design and Methods:** A secondary analysis of the Overdose Response and Take Home Naloxone (ORTHN) project, comparing participant characteristics, and changes in knowledge, attitudes and self-efficacy regarding overdose and response, following a brief THN intervention in participants who had previously undertaken THN interventions, compared to those with no prior THN interventions. Data was analysed for those completing both baseline (pre-ORTHN intervention) and follow up (3-months later).

**Results:** Ninety-four participants completed both research interviews, of whom 29 (31%) had previously completed a THN intervention. There were few differences in baseline demographics or overdose histories between the two groups. Knowledge regarding overdose response and naloxone use indicated high baseline levels in both groups. Those with no prior THN training had lower rates of self-efficacy at baseline, and significantly improved following the ORTHN intervention, whereas those with prior THN training had higher baseline levels of self-efficacy, which was maintained following the intervention.

**Discussion and Conclusions:** Individuals who have previously undertaken a THN intervention may not require repeat training when renewing naloxone supplies, increasing the efficiency of THN interventions.

## Introduction

Marked increases in opioid overdoses internationally in the past decade has seen the proliferation of take-home-naloxone (THN) interventions, including provision of naloxone, a short-acting opioid antagonist to people who use opioids and their carers [1, 2]. Most THN interventions involve training of recipients. The European Monitoring Centre for Drugs and Drug Addiction describes training as “an essential part of take-home naloxone distribution programmes that can effectively increase participants’ knowledge, confidence and skills in managing an opioid overdose” (p. 8) [2]. Common themes include signs and risk factors of an opioid overdose, actions to take (e.g., rescue breathing, naloxone administration), and aftercare procedures [3-8].

THN programs have operated in Australia since 2012 [1], with initial programs utilising an advanced, lengthier approach [in line with 2], and later programs using briefer training in efforts to increase THN dissemination [9]. Evaluation of brief Australian THN interventions [9] indicate they adequately enhance knowledge, attitudes and self-efficacy. Training remains a standard recommendation of Australian THN interventions, embedded in many organisational procedures.

Changes in the administration of the opioid treatment program in Australia such as reduced contact and increased takeaways due to the COVID19 pandemic has created greater urgency in the need to distribute THN given the potential increased risks for opioid overdose, necessitating consideration of how to streamline THN dissemination. Training clients requires a skilled workforce and staff time, which may be currently limited. One approach to reduce barriers to THN access could be to remove the requirement for retraining consumers upon resupply of THN. Whilst there are international examples of programs where training of consumers is not required [10], these do not appear to have been evaluated. One retrospective cohort study identified no significant differences in overdose responses between trained and untrained rescuers [11], suggesting the benefits of training may be overstated.

We aim to examine if repeating training to those clients with prior THN intervention is necessary. This is a secondary analysis of a previously reported study examining a brief THN intervention targeting people at risk of overdose, delivered in Alcohol and other Drug (AoD), needle syringe programs (NSP), and a supervised injecting facility in New South Wales, Australia [12]. The evaluation of the original ORTHN (Opioid Response and Take Home Naloxone) project demonstrated that THN interventions improved knowledge, attitudes and self-efficacy in participants, with assessment immediately before and three months after the intervention. Here, we examine whether these improvements differ for prior THN participants compared to those with no prior THN exposure.

## Methods

### Design and settings

Details of the ORTHN program and evaluation have been previously described [12]. In short, psychoeducation and free naloxone was provided by a broad range of health workers (nurses, counsellors, AoD and NSP workers) in a 10-30 minute brief intervention to over 600 clients. We recruited 145 participants to participate in research interviews immediately before the ORTHN intervention, with a 3-month follow-up interview. Prior to the implementation of the ORTHN program, THN interventions had been delivered in the preceding 2 years at various sites that participated in the ORTHN evaluation, such that some participants had prior THN intervention exposure. The project was approved by South East Sydney Local Health District (SESLHD) Human Research Ethics Committee as a multisite study with local research governance approvals.

### Measures

#### Demographic questions

Age, gender, employment, relationship and educational status.

#### Overdose-related knowledge, attitudes, and ratings of self-efficacy

Overdose knowledge was assessed using a modified Opioid Overdose Knowledge Scale (OOKS) [13] with four sub-scales: (overdose risk factor, signs, responses, and naloxone use; see [12] for more information). A modified Opioid Overdose Attitudes Scale (OOAS) [13] included five questions, described in the Results; the five-point Likert responses (strongly disagree to strongly agree) were modified to a binary outcome of ‘endorsed’ (agree/strongly agree) and ‘not endorsed’ (the rest).

#### Previous experiences with Naloxone

A Yes or No response to “have you ever been trained in how to use naloxone?”

### Statistical analysis

Chi-squared analyses (or Fisher’s Exact tests where appropriate) and Independent samples *t*-tests compared differences in demographics and overdose history between those with and without prior ORTHN naloxone training. Independent samples *t*-tests and repeated measures tests were conducted at baseline and at follow-up to compare those with and without prior naloxone training regarding knowledge (OOKS). Logistic mixed-effects models for repeated measures (MMRM) regressions were used to analyse each of the five binary attitudes (proportion ‘endorsed’ vs ‘not endorsed’). Prior training, a two-level categorical variable (prior vs no prior THN) was the between-subjects fixed predictor in the model, Time the two-level categorical within-subjects predictor (baseline vs follow up), and participant ID the random effect. Within these models the simple effects of time at each level of prior THN were tested, as well as the interaction between prior THN and time. In the event that any MMRM failed to converge (for example if either 0% or 100% of one group endorsed the attitude at both baseline and follow-up) a two-sample test of equality of proportions was performed, testing the difference between the proportion who endorsed the attitude in the prior and no-prior THN groups, at both baseline and follow-up. Although these tests do not account for repeated measures, and hence are not as accurate/reliable as an MMRM, in combination they describe the between-group difference in amount of change in proportion from baseline to follow-up.

## Results

### Participants

Baseline and follow-up questionnaires were completed by 94 participants and are included in subsequent analyses. Most (65/94, 69%) had no prior THN intervention before the ORTHN project (Table 1). A significantly greater proportion of the prior THN group had completed tertiary education, had witnessed an OD in their lifetime, and had previously administered naloxone to someone.

**Table 1:** Participant demographic and overdose history characteristics.

| Variable | Prior THN<br>(n=29) | No Prior<br>THN<br>(n=65) | TOTAL<br>N=94 | Statistical<br>Test | p |
| --- | --- | --- | --- | --- | --- |
| Age (years), Mean±SD | 42.3±9.5 | 41.1± 9.1 | 41.5±9.1 | $t(92)=-0.57$ | 0.57 |
| Gender, n (%) |  |  |  |  |  |
| Male | 18 (62.1%) | 38 (58.5%) | 56 (59.6%) | $\chi^2(1,94) = 0.11$ | 0.82 |
| Female | 11 (37.9%) | 27 (41.5%) | 38 (40.4%) |  |  |
| Highest educational<br>qualification, n (%) |  |  |  |  |  |
| High school | 20 (69.0%) | 57(87.7%) | 77 (81.9%) | FET = 5.81 | 0.03 |
| Tafe/University | 9 (31.0%) | 7 (10.8%) | 16 (17.0%) |  |  |
| Age first injected drugs<br>Mean±SD | 17.4± 4.4 | 19.1± 5.0 | 18.6±4.9 | $t(91) = 1.62$ | 0.58 |
| Participants who have ever<br>witnessed an overdose, n (%) | 29 (100.0%) | 54 (83.1%) | 83 (88.3%) | FET = NA | 0.02<sup>a</sup> |
| Participants who have<br>witnessed an OD in last 3<br>months, n (%) | 9 (39.1%) | 14 (25.9%) | 23 (27.7%) | $\chi(1,83) = 0.25$ | 0.80 |
| Participants with previous<br>overdose (lifetime), n (%) | 15 (51.7%) | 39 (60.0%) | 54 (57.4%) | $\chi(1,94) = 0.56$ | 0.50 |
| Age first OD, Mean±SD | 25.8±12.5 | 23.8± 6.6 | 24.4±8.5 | $t(52) = -0.76$ | 0.45 |
| Participants overdosed in last<br>3 months, n (%) | 1/15 (6.7%) | 2/39 (5.1%) | 3/54 (5.6%) | FET = NA | 1.00 |
| Have you ever given naloxone<br>to anyone?, n (%) |  |  |  |  |  |
| Yes | 12 (41.4%) | 4 (6.2%) | 16 (17.0%) | FET = NA | <0.001 |
| No | 17 (58.6%) | 61 (93.8%) | 79 (83.%) |  |  |

FET = Fisher's exact test

### Knowledge regarding overdose and naloxone

There were no significant differences between the groups at baseline, no significant changes over time (baseline to follow-up), and no significant time by training interaction effects (i.e., prior THN status did not significantly impact this result), all *p*s > 0.10.

### Attitudes and self-efficacy regarding overdose and naloxone

Responses in each group and at each time point are presented in Figure 1.

**Figure 1:**
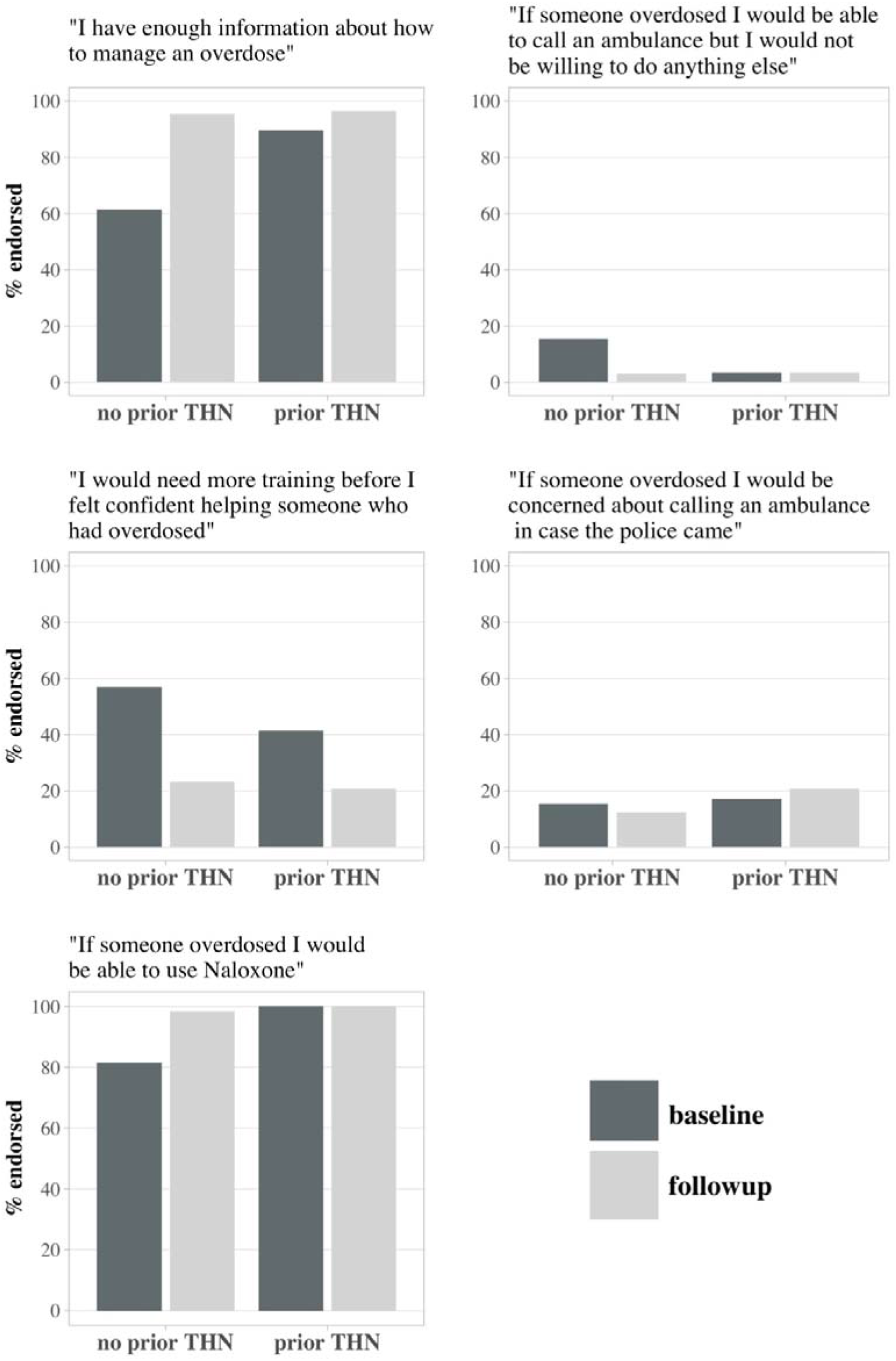
A comparison of overdose and naloxone attitudes for those with prior and no-prior THN intervention expsoure, at baseline and follow up.

Four of the five logistic MMRMs converged. There was no significant interaction between training and time in any of these models (*p-*range 0.252 – 0.450), however, there were consistent patterns across variables in the simple effects of time at each level of prior THN exposure. Among participants with no prior THN, the ORTHN training was associated with a significant increase in the odds of endorsing the statement “I have enough information about how to manage an overdose” (OR=12.9, 95%CI=3.6, 47.0, *p*<0.001), whereas there was no significant increase among the group with prior THN intervention (OR=3.2, 95%CI=0.4, 29.1, *p*=0.298). There was a significant reduction after ORTHN intervention in endorsement of the statement “If someone overdosed I would be able to call an ambulance but I would not be willing to do anything else”, in those with no prior THN (OR=0.2, 95%CI=0.0, 0.8, *p*<0.001), but no significant change in those with prior THN, (OR=1.0, 95% CI=0.1, 14.5, *p*=0.999). Both groups had reductions in the odds of endorsing the statement “I would need more training before I felt confident helping someone who had overdosed” after ORTHN training, (OR=0.2, 95%CI=0.1, 0.5, *p*<0.001 in ‘no prior THN’; and OR=0.4, 95%CI=0.1, 1.1, *p*=0.064 in prior THN). There were no significant reductions in the odds of endorsing “If someone overdosed I would be concerned about calling an ambulance in case the police came” for either group. The MMRM based on the statement “If someone overdosed I would be able to use Naloxone” failed to converge due to the fact all participants in the group with prior THN training endorsed the statement at both time points. The two-sample test for equality of proportions revealed that at baseline a significantly lower proportion of participants in the group with no prior THN training endorsed the statement than in the group who had some THN training, *χ*^2^(1)=4.6, *p*=0.032, but this between-group difference disappeared at follow-up *χ*^2^ (1)=0.0, *p*=1.000.

## Discussion

The findings indicate that those with no prior THN intervention significantly improved their attitudes and self-efficacy following the ORTHN intervention – highlighting the importance of training in the initial THN interventions with a client. These concepts appear to be important in an individual’s response to overdose and the use of naloxone [14]. In contrast, those with prior THN interventions had moderate to high levels of self-efficacy prior to the current ORTHN intervention, with minimal change afterwards – suggesting little benefit from ‘repeat training’. Furthermore, levels of knowledge regarding overdose and naloxone use were high in both groups prior to THN interventions, with little room for improvement – a finding consistent with other Australian evaluations [3].

Our findings suggest that the delivery of THN interventions can be adapted to not require training in people who have previously received a THN intervention. Further, it has been established that knowledge around naloxone is retained well over time [6]. People taking part in THN interventions should be asked whether they have previously had an intervention, and have sufficient knowledge and confidence to respond to a suspected overdose and use naloxone, and if so, then naloxone should be supplied without repeat training. This is relevant given the increasing emphasis upon THN interventions being delivered in community pharmacies, with inconsistent pharmacist training and stigma concerns have been identified [15].

The main study limitation is the sample size – with only 29 of the 94 participants having had prior THN interventions, prohibiting meaningful examination of actual overdose responses, nor identification of subgroups who may benefit from repeat training (e.g. cognition problems).

In conclusion, these findings suggests that repeat THN interventions can focus on supply of naloxone, with less emphasis upon the training components of conventional THN interventions.

## Data Availability

Available upon request

## Declarations

### Authors’ contributions

N.L. designed the study, provided oversight of study conduct, and led the writing of the manuscript. L.M. contributed to the design of the study, supervised data collection, and contributed to analyses and manuscript writing. M.B. was responsible for data collection and manuscript writing. L.Mills and C.R contributed to analysis. S.N., M.E.H., and P.R. contributed to study design and project oversight. All authors supported the conduct of the project and contributed to and approved the final manuscript.

## Acknowledgements

In particular, the authors would like to thank Professor Paul Dietze for his extensive and valuable guidance on the direction of the manuscript. The authors wish to acknowledge the support and participation of clients and staff in participating services, including peer researchers at NUAA. The authors also acknowledge the assistance and support of NSW Health, particularly the Offices of the Chief Pharmacist and Chief Medical Officer, and support from AoD Branch within the MoH. The project was funded by a grant from the NSW Health Translational Research Grant Scheme, and with funding support from University of Sydney. Study medication (Prenoxad®) was purchased from Phebra.

## References

[1] Dwyer R, Olsen A, Fowlie C, Gough C, van Beek I, Jauncey M, et al. An overview of take□home naloxone programs in Australia. Drug Alcohol Rev. 2018;37(4):440–9.

[2] Strang JS, McDonald R. Preventing opioid overdose deaths with take-home naloxone. In: European Monitoring Centre for Drugs and Drug Addiction, editor. Luxembourg: Publications Office of the European Union; 2016.

[3] Dietze PM, Draper B, Olsen A, Chronister KJ, van Beek I, Lintzeris N, et al. Does training people to administer take□home naloxone increase their knowledge? Evidence from Australian programs. Drug Alcohol Rev. 2018;37(4):472–9.

[4] McAuley A, Lindsay G, Woods M, Louttit D. Responsible management and use of a personal take-home naloxone supply: a pilot project. Drugs (Abingdon Engl). 2010;17(4):388–99.

[5] Seal KH, Thawley R, Gee L, Bamberger J, Kral AH, Ciccarone D, et al. Naloxone distribution and cardiopulmonary resuscitation training for injection drug users to prevent heroin overdose death: a pilot intervention study. J Urban Health. 2005;82(2):303–11.

[6] Strang J, Manning V, Mayet S, Best D, Titherington E, Santana L, et al. Overdose training and take□home naloxone for opiate users: prospective cohort study of impact on knowledge and attitudes and subsequent management of overdoses. Addiction. 2008;103(10):1648–57.

[7] Wagner KD, Valente TW, Casanova M, Partovi SM, Mendenhall BM, Hundley JH, et al. Evaluation of an overdose prevention and response training programme for injection drug users in the Skid Row area of Los Angeles, CA. Int J Drug Policy 2010;21(3):186–93.

[8] Lintzeris N, Monds LA, Bravo M. The NSW Overdose Response with Take Home Naloxone (ORTHN) Project: Establishing a model of care for the delivery of ORTHN interventions for clients attending AoD, NSP and related outreach settings. In: Service SESLHDDaA, editor. Sydney 2019.

[9] Behar E, Santos G-M, Wheeler E, Rowe C, Coffin PO. Brief overdose education is sufficient for naloxone distribution to opioid users. Drug and alcohol dependence. 2015;148:209–12.

[10] Petty J, McNally S, Ryan J. Saving lives: Australian naloxone access model Melbourne: Penington Institute 2018.

[11] Doe-Simkins M, Quinn E, Xuan Z, Sorensen-Alawad A, Hackman H, Ozonoff A, et al. Overdose rescues by trained and untrained participants and change in opioid use among substance-using participants in overdose education and naloxone distribution programs: a retrospective cohort study. BMC Public Health. 2014;14(1):297.

[12] Lintzeris N, Monds LA, Bravo M, Read P, Harrod ME, Gilliver R, et al. Designing, implementing and evaluating the overdose response with take□home naloxone model of care: An evaluation of client outcomes and perspectives. Drug Alcohol Rev. 2020;39(1):55–65.

[13] Williams AV, Strang J, Marsden J. Development of Opioid Overdose Knowledge (OOKS) and Attitudes (OOAS) Scales for take-home naloxone training evaluation. Drug Alcohol Depend. 2013;132(1-2):383–6.

[14] Tormohlen KN, Tobin KE, Davey-Rothwell MA, Latkin C. Low overdose responding self-efficacy among adults who report lifetime opioid use. Drug Alcohol Depend. 2019;201:142–6.

[15] Olsen A, Lawton B, Dwyer R, Taing M-W, Chun KLJ, Hollingworth S, et al. Why aren’t Australian pharmacists supplying naloxone? Findings from a qualitative study. International Journal of Drug Policy. 2019;69:46–52.

